# Fall experiences of ambulatory children and adults with cerebral palsy: a qualitative analysis

**DOI:** 10.1101/2025.03.22.25324449

**Authors:** Marissa Esterley, Linda E. Krach, Kari Pederson, Sandy Callen Tierney, Nathan G. Wandersee, Elizabeth R. Boyer, Cerebral Palsy Research Network

**Author notes:** Corresponding Author: Elizabeth R. Boyer, PhD, Gillette Children’s Specialty Healthcare 200 University Avenue East, Saint Paul, MN 55101, USA.

## Abstract

**Aim:** To qualitatively assess causes, adaptations, and psychosocial impact of falls, and solutions for safer environments as shared by persons diagnosed with cerebral palsy (CP).

**Method:** Ambulatory adults with CP (n=165) and caregivers of ambulatory children with CP (n=151) responded to four open-ended falls questions. Deductive and inductive content analysis was conducted. **Results:** Eight themes emerged (psychological, physical, avoid, adapt, people, environment, policy, healthcare). Participants elaborated on fall causes (aging, physical, mental, environmental, and situational), mechanics (most often trips), repercussions (psychological and physical), adaptations, difficulty getting up, and aspirations for themselves and society. Caregivers and adults detailed various adaptations to or deliberate avoidance of high-risk situations (e.g. uneven surfaces, crowds). Specific suggestions for environmental accessibility (e.g. more handrails), societal behavioral responses (give autonomy, be patient), healthcare practice, and policy were made.

**Interpretation:** This study offers profound insights into how individuals with CP navigate the challenges of falls and how people and surroundings both positively and negatively affect their fall-related experiences. Many issues identified were multifactorial, requiring multidimensional, non-ableist solutions. Thus, the onus to address these issues is shared. Participants offered simple, but impactful, actions that could be taken immediately to support the creation of safer physical and psychological environments.

**WHAT THIS PAPER ADDS:** 1. People should ask *if* and *how* to help when someone falls.

2. Falls trigger anxiety, embarrassment, and avoidance, often outweighing physical injuries’ impact.

3. Participation can be enhanced through more inclusive activities and environments.

4. Safe falling strategies should be taught at all ages.

5. Multifaceted solutions include conversations regarding falls with clinicians and addressing policy shortcomings.

---

Ambulatory individuals with cerebral palsy (CP) often experience restricted mobility, increased unsteadiness, and elevated risk and concern for falling.^1–3^ Falls can cause embarrassment, activity avoidance, and injury.^3–8^ The negative emotional and behavioral responses may become more prominent in adolescents,^8^ though high levels have been reported through middle adulthood.^3^ Over half of ambulatory children and adults with CP fall at least once per year,^3,6,9,10^ which is two to three times higher than the general older adult population.^11^ Consequently, individuals with CP experience the physical and psychological sequelae of falls throughout their lives. However, there is a dearth of literature on the lived experiences related to falls, despite the higher risk of falls.^7,12^

Understanding CP and falls requires a multidimensional framework that considers the biological, psychological, and societal contributors to disability and health.^13^ A small study reflected this by identifying helpful and unhelpful attitudes, practices, and environment modifications that affect participation in the face of fall risk.^8^ Avoiding activities and seeking support were common and sometimes beneficial,^5,8^ but they can lead to isolation, reduced independence, and stigmatization, especially by some healthcare professionals’ limited awareness of CP-specific challenges.^14^ Conversely, when healthcare professionals understand their patients’ daily lives, positive outcomes can result.^14^ Increasing awareness of the lived experience surrounding falls will inform resource allocation, clinical services, and research. Hence, our study purpose was to describe the experiences of individuals with CP related to falls and their repercussions and how clinicians and society can help mitigate the impact of falls across the lifespan.

## METHODS

The University of Minnesota Institutional Review Board approved this study, which was part of a larger mixed methods falls study.^3^ Qualitative findings are reported here. The inclusion criteria were individuals with CP, aged 5-89, Gross Motor Function Classification System (GMFCS) levels I-III, who read English. Acknowledgement of consent was obtained from caregivers for minors (<18 years) and self for adults (≥18 years).

Recruitment occurred from January to October 2022 via emails from individuals seen at Gillette Children’s from 2011-2021, social media listings in 2022, and emailed surveys to adults within the Cerebral Palsy Research Network’s Community Registry.

REDCap (Research Electronic Data Capture) was used for screening, electronic consent, and survey administration. Caregivers were asked to consult their child, as appropriate. Four optional open-ended falls questions based on the literature^8^ and conversations with adults with CP were created: 1) Feel free to share any more information or provide clarifications regarding the above questionnaire on consequences of falling, 2) Describe how you may have adapted to perform certain activities to avoid falls, 3) Provide clarifications regarding the above questionnaire on balance confidence and activity avoidance, and 4) Provide any other comments about balance, trips, or falls, including what clinicians, families, or society can do differently to make physical and social environments safer or decrease your concerns about falls.

### Data Analysis

Child/caregiver and adult responses were analyzed together. We used Framework Method with deductive approaches guided by our research questions and inductive approaches guided by data content analysis.^15^ Responses were independently analyzed by two authors (ME, ERB).

During familiarization, the textual data were coded, codes were organized into potential themes and subthemes, maintaining alignment with the four open-ended research questions. A working codebook was created, and researchers defined the central themes and subthemes. A thematic framework matrix was created in an electronic spreadsheet where columns represented subthemes, and rows represented participants. Throughout the charting phase, sentences, phrases, and paragraphs of responses were labeled in the framework matrix. People with lived experience (KP, ST, NW) informed the codebook and joined disagreement discussions to help reach consensus. After further review, subthemes were finalized to best represent the original responses.

## RESULTS

The final sample in the larger study included 381 participants, of which 316 provided at least one response to the four optional open-ended questions (Table 1).

**Table 1.** Demographics.

|  | **All** | **Children** | **Adults** |
| --- | --- | --- | --- |
| **n** | 316 | 151 | 165 |
| **Age (y)**, median<br>(25%tile, 75%tile) min-max] | 19 (10,31) [5-76] | 10 (7,14) [5-17] | 30 (25,50) [18-76] |
| **Gender**, n (%) |  |  |  |
| **female** | 165 (52%) | 64 (44%) | 101 (63%) |
| **male** | 142 (45%) | 83 (56%) | 59 (37%) |
| **Non-binary/not specified** | 9 (3%) | 4 (3%) | 5 (3%) |
| **GMFCS level**, n (%) |  |  |  |
| **I** | 98 (31%) | 49 (32%) | 49 (30%) |
| **II** | 142 (45%) | 66 (44%) | 76 (46%) |
| **III** | 76 (24%) | 36 (24%) | 40 (24%) |
| **Topography**, n (%) |  |  |  |
| **Not specified** | 7 (2%) | 5 (3%) | 2 (1%) |
| **Monoplegia** | 20 (6%) | 11 (7%) | 9 (6%) |
| **Hemiplegia** | 94 (30%) | 53 (35%) | 41 (25%) |
| **Diplegia** | 97 (31%) | 37 (25%) | 60 (37%) |
| **Triplegia** | 30 (10%) | 21 (14%) | 9 (6%) |
| **Quadriplegia** | 64 (21%) | 23 (15%) | 41 (25%) |

Across all four questions, two central themes—internal and external—and eight subthemes emerged (internal: mental, physical, avoid, adapt; external: people, environment, policy, healthcare). Participants elaborated on fall causes, mechanics, repercussions, frustrations, and desired changes for themselves and society. Responses to the second question were divided into five additional subthemes related to avoid (public, physical exertion, terrain) and adapt (physical behaviors, mental). Internal themes are discussed first (Table 2) followed by external themes (Table 3).

**Table 2.** Example quotes from central theme 1, internal, defined as thoughts, actions or characteristics of the individual with CP.

|  |
| --- |
| **MENTAL:** Mental ideas, thoughts, feelings, or emotions that an individual with CP can be both causes and effects or consequences of a fall |
| *"I am more concerned about the embarrassment of falling rather than the potential physical damage." [Q1] [20-29y, GMFCS II, triplegia]* |
| *"...mental fatigue/being distracted is the cause of the overwhelming majority of my falls. If I'm not THINKING enough to pick up my foot, BAM." [Q1] [40-49y, GMFCS II, diplegia]* |
| *"Sometimes I worry more about their reaction before even thinking about any pain. I worry people will see my falls and worry I shouldn't be independent. I worry they will stop seeing me as an equal and more as a liability." [Q1] [30-39y, GMFCS II, quadriplegia]* |
| *"I have a lot of anxiety about falling. I've had panic attacks before and after falling. For years I refused to use an assistive device." [Q1] [40-49y, GMFCS III, quadriplegia]* |
| *"I think fear and anxiety around falling contribute a lot to my inability to balance when performing some of the activities...The older I get, the less confident I am in my balance and the slower my reaction times become." [Q4] [30-39y, GMFCS II, diplegia]* |
| **PHYSICAL:** The individual with CP's physical impairments that may cause a fall, or physical/bodily consequences of falling |
| *"When [my child] falls it is often due to more than one variable. Example uneven surface and muscle fatigue. He often gets very angry and sad when he falls and lashes out." [Q1] [caregiver of 0-9y, GMFCS II, quadriplegia]* |
| *"Often fall as a startle reflex." [Q1] [30-39y, GMFCS I, diplegia]* |
| *"I wish it wasn't so hard for me to get up from the floor without assistance. That is why I am so fearful of falling." [Q1] [50-59y, GMFCS III, diplegia]* |
| *"If I get nervous then my muscles tense up, and it takes a bit for them to calm down." [Q4] [30-39y, GMFCS I, quadriplegia]* |
| *"If I am tired or in a hurry, I am more likely to fall." [Q4] [50-59y, GMFCS II, diplegia]* |
| *"As a senior falls are more fearful now. That fear will also tightens the body up which increases the likelihood of falling. You Worry more about injuries. As a younger person you got up quickly so no one would see you, shook it off and went on. Now if I fall I may need help to get up or have to crawl to some piece of furniture in order to get up. Decreases independence." [50-59y, GMFCS II, diplegia]* |
| **AVOID:** Activities or spaces intentionally not performed/attended by the individual with CP due to their real or perceived fall risk and/or any physical or mental repercussions if a fall occurs |
| *"It lowers confidence, increases fear and does not want to go out and access life as a result. Especially after a hard fall." [Q1] [caregiver of 10-19y, GMFCS III, quadriplegia]* |
| *"I have fairly significant anxiety in large crowds as a direct result of all the falls I've experienced. (And the subsequent fear of falling or being knocked over.)" [Q2] [30-39y, GMFCS III, quadriplegia]* |
| *"Sometimes I just sit there and watch everyone else have fun, because I hate my disease." [Q2] [caregiver of 10-19y, GMFCS II, diplegia]* |
| *"[My child] will avoid sporting activities and other youth events because of the possibility he could fall. He's quit soccer because of this, dreads school track and field day, etc." [Q2] [caregiver of 0-9y, GMFCS II, diplegia]* |
| *"If there are a set of stairs/slope, I will absolutely find another route." [Q2] [30-39y, GMFCS I, hemiplegia]* |
| **ADAPT:** Adaptations (physical, mental or medical treatments) by individuals with CP due to their real or perceived fall risk and/or any physical or mental repercussions if a fall occurs |
| *"Fortunately I learned how to take a fall so that I can hopefully prevent major injuries." [Q1] [60-69y, GMFCS II, hemiplegia]* |
| *"Usually if he feels unbalanced, he will voluntarily fall, so the fall hurts less. I would say most of his falls are because he feels like the fall is coming, so instead of trying to catch or correct himself, he chooses to fall at his own pace." [Q1] [caregiver of 10-19y, GMFCS II, triplegia]* |
| *"I often times use my walker, brace myself against something or hold on to something while performing an activity where falling is a concern." [Q2] [20-29y, GMFCS III, quadriplegia]* |
| *"I will psych myself up to do them. If I know it is something that I know there is no way I can do it, I will just automatically ask for help or not do it." [Q2] [30-39y, GMFCS I, quadriplegia]* |
| *"I try to do risky things with optimal mental awareness and as slowly as possible in case I have to change my approach or adaptation mid-stream. Optimal mental acuity and muscle strength/tone is key for me; which is one reason I stay away from muscle relaxants. I live alone; so if I loose[sic] my ability to anticipate things and think things through in risky or potentially risky situations, I'm screwed. However for me, now that I am older, most of the danger in falling is the difficulty/ expenditure of energy and time in getting up off the floor." [Q2] [50-59y, GMFCS III, quadriplegia]* |

Q1: Question 1, Q2: Question 2, Q3: Question 3, Q4: Question 4

**Table 3.**
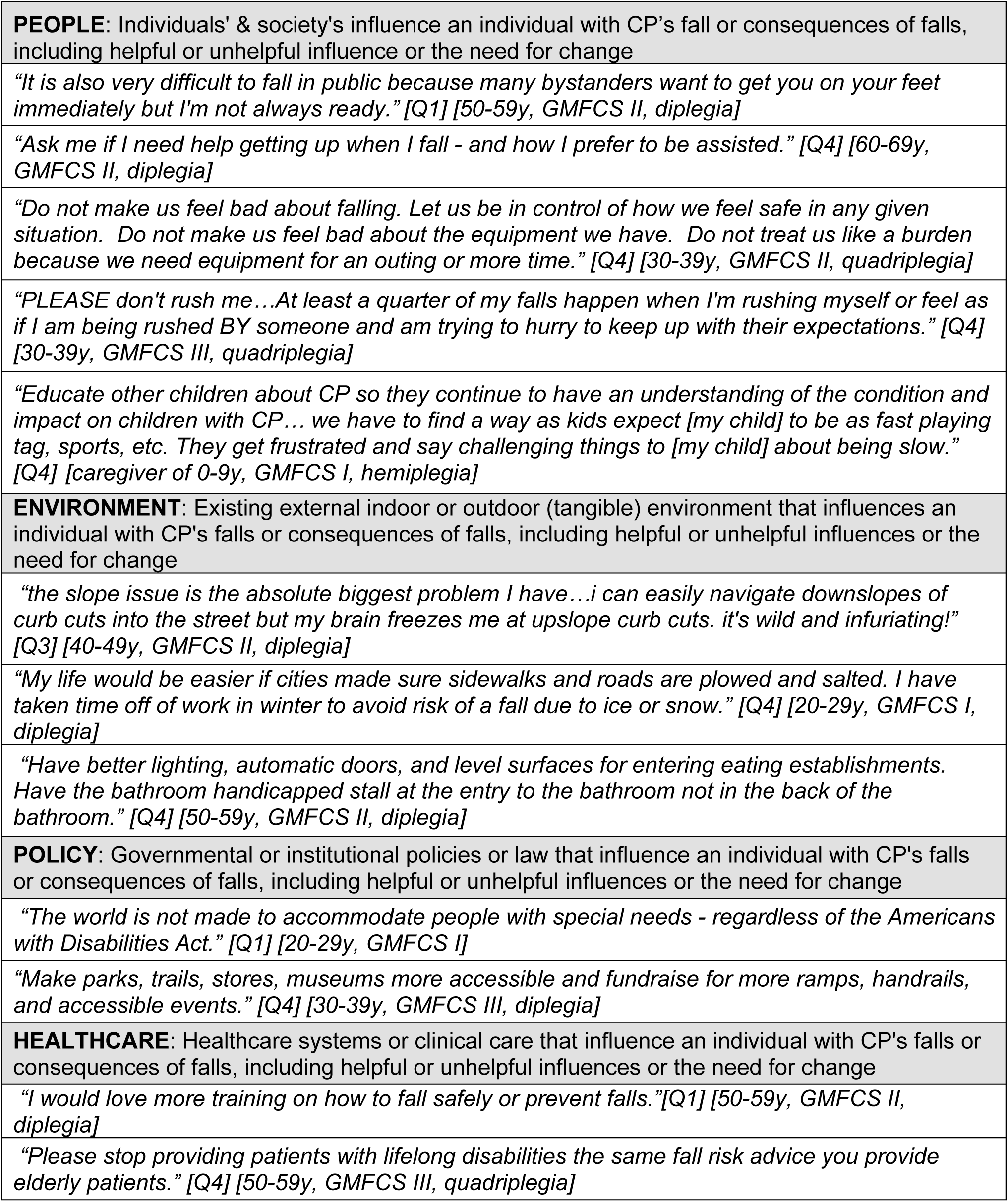
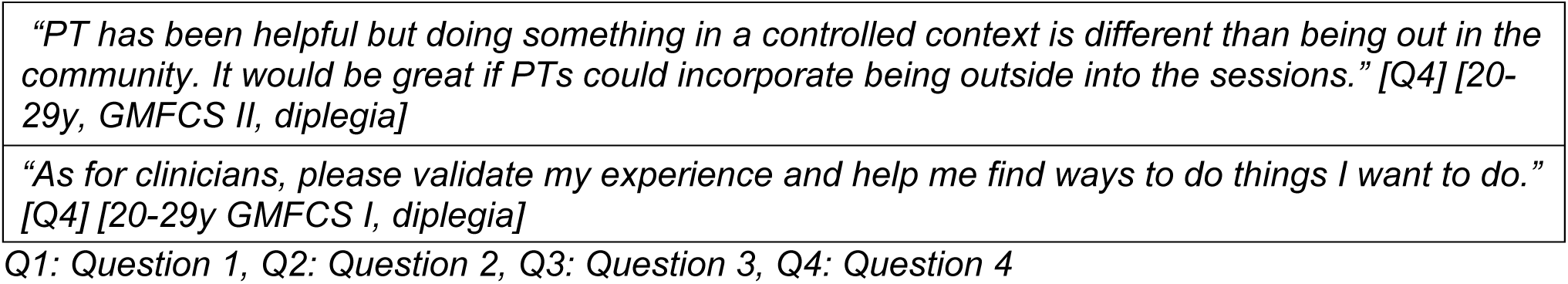
Example quotes from central theme 2, external, defined as thoughts, actions or things external to the individual with CP.

|  |
| --- |
| **PEOPLE:** Individuals' & society's influence an individual with CP's fall or consequences of falls, including helpful or unhelpful influence or the need for change |
| *"It is also very difficult to fall in public because many bystanders want to get you on your feet immediately but I'm not always ready."* [Q1] [50-59y, GMFCS II, diplegia] |
| *"Ask me if I need help getting up when I fall - and how I prefer to be assisted."* [Q4] [60-69y, GMFCS II, diplegia] |
| *"Do not make us feel bad about falling. Let us be in control of how we feel safe in any given situation. Do not make us feel bad about the equipment we have. Do not treat us like a burden because we need equipment for an outing or more time."* [Q4] [30-39y, GMFCS II, quadriplegia] |
| *"PLEASE don't rush me...At least a quarter of my falls happen when I'm rushing myself or feel as if I am being rushed BY someone and am trying to hurry to keep up with their expectations."* [Q4] [30-39y, GMFCS III, quadriplegia] |
| *"Educate other children about CP so they continue to have an understanding of the condition and impact on children with CP... we have to find a way as kids expect [my child] to be as fast playing tag, sports, etc. They get frustrated and say challenging things to [my child] about being slow."* [Q4] [caregiver of 0-9y, GMFCS I, hemiplegia] |
| **ENVIRONMENT:** Existing external indoor or outdoor (tangible) environment that influences an individual with CP's falls or consequences of falls, including helpful or unhelpful influences or the need for change |
| *"the slope issue is the absolute biggest problem I have...i can easily navigate downslopes of curb cuts into the street but my brain freezes me at upslope curb cuts. it's wild and infuriating!"* [Q3] [40-49y, GMFCS II, diplegia] |
| *"My life would be easier if cities made sure sidewalks and roads are plowed and salted. I have taken time off of work in winter to avoid risk of a fall due to ice or snow."* [Q4] [20-29y, GMFCS I, diplegia] |
| *"Have better lighting, automatic doors, and level surfaces for entering eating establishments. Have the bathroom handicapped stall at the entry to the bathroom not in the back of the bathroom."* [Q4] [50-59y, GMFCS II, diplegia] |
| **POLICY:** Governmental or institutional policies or law that influence an individual with CP's falls or consequences of falls, including helpful or unhelpful influences or the need for change |
| *"The world is not made to accommodate people with special needs - regardless of the Americans with Disabilities Act."* [Q1] [20-29y, GMFCS I] |
| *"Make parks, trails, stores, museums more accessible and fundraise for more ramps, handrails, and accessible events."* [Q4] [30-39y, GMFCS III, diplegia] |
| **HEALTHCARE:** Healthcare systems or clinical care that influence an individual with CP's falls or consequences of falls, including helpful or unhelpful influences or the need for change |
| *"I would love more training on how to fall safely or prevent falls."*[Q1] [50-59y, GMFCS II, diplegia] |
| *"Please stop providing patients with lifelong disabilities the same fall risk advice you provide elderly patients."* [Q4] [50-59y, GMFCS III, quadriplegia] |

*"PT has been helpful but doing something in a controlled context is different than being out in the community. It would be great if PTs could incorporate being outside into the sessions." [Q4] [20-29y, GMFCS II, diplegia]*
*"As for clinicians, please validate my experience and help me find ways to do things I want to do." [Q4] [20-29y GMFCS I, diplegia]*
Q1: Question 1, Q2: Question 2, Q3: Question 3, Q4: Question 4

### Psychological

Multiple participants identified as “expert fallers,” stating falls are a part of their lives and require acceptance, adaptation, and resilience. Psychological causes of falls reported were mental fatigue, distraction, and overthinking. Most responses centered on psychosocial repercussions, highlighting how perspectives like resilience or despondence can impact participation and quality of life. Participants struggled with embarrassment, anxiety, panic attacks, and insecurity; these affected their balance and were more worrisome than the physical ramifications of falls. A caregiver mentioned their child felt like other children noticed the physical difficulty of some tasks, increasing the child’s fear and insecurity. Others were resilient after falls. Caregivers wanted their child to feel empowered, not embarrassed, when asking for help.

### Physical

Participants described physical impairments contributing to falls, including fatigue, weakness, tight muscles, low tone, poor balance, and age-related changes. Trips caused by foot clearance issues were the predominant cause of falls rather than slips. Some reported being startled, especially in public, as a substantial contributor to falls, prompting them to sometimes avoid public spaces. Children and adults alluded to excitement or nervousness causing muscle tension, which caused falls. Participants suspected comorbidities may also cause falls, like autism, seizures, cerebral/cortical visual impairment (CVI: visual impairment due to damage to visual processing rather than eye structures), blindness, and cervical stenosis. The effects of falls included pain, minor injuries, and major injuries like fractures and concussions. Physical repercussions were reported to change over time, especially difficulty getting up.

### Avoid

Participants described how concern about falling directly affected their participation. Many avoid crowds and public spaces. One participant, in middle adulthood, worried about falling and so avoids high-risk situations since an injury could impede their ability to provide for their family as the primary household earner. Participants avoided uneven, irregular, slick, or unknown terrain. One participant took time off work to avoid walking in wintry conditions. Others discussed taking longer routes to avoid tenuous areas, staying on the perimeter of crowds and near walls for support. Avoidance of sports and physical activity was common for children and adults.

### Adapt

Participants discussed physical and mental adaptations to mitigate fall risk, which were more common than avoidance examples. Physical adaptations included sitting to dress and shower, crawling/scooting, wearing more supportive shoes, strength and balance exercises, and using assistive devices, braces/orthoses, external structures (e.g., furniture, walls), or people for support. Scooting and crawling were reported by children and adults in GMFCS levels II and III. Some described using assistive devices (e.g., walker) to create space around them. Other physical adaptations included using hiking sticks and looking down. Mental adaptations included hypervigilance and planning when going out in public. Participants noted the importance of concentrating on walking and paying attention to surfaces, objects, or crowds that might increase fall risk. Other common themes included anticipation, understanding one’s limitations, and learning when to seek help.

### People

Participants discussed how others helped or harmed their fall experiences. A companion’s presence or assistance for safety during daily activities (e.g., showering, and especially reaching for items on the floor) was reported by many as helpful. Participants discussed frustrations and desires to change others’ reactions and behaviors; strangers can cause a fall or complicate the physical or mental recovery. People often felt crowded by strangers or rushed to move quickly. Asking people to remove shoes upon entering a house was shared as an example of ableism. Many experienced unwanted attention after falls in public. Participants urged strangers witnessing a fall to avoid overreacting, instead, ask if assistance is needed, and then ask *how* to help. A caregiver desired resources for their child’s school to help educators understand, prevent, and respond to falls.

### Environment

The environment undeniably contributed to falls. Trip hazards included rugs, toys, uneven surfaces (e.g., curbs, stairs), and crowds. Familiarity affected fall risk and concern; stairs in one’s home or school were concerning, but stairs in unfamiliar spaces caused angst.

Participants criticized misplaced or inadequate accessibility features. Numerous participants experienced anxiety on downward slopes, especially steeper slopes or those without handrails. Participants offered simple suggestions to improve accessibility, such as clearing public walkways (e.g., shopping aisles, sidewalks). And while some wanted more slopes, others wanted less, instead preferring low incline, deeper stairs. Decorations wrapped around handrails were criticized for obstructing use. One participant discussed the dangers of being outdoors in warmer climates, because falls on the ground can cause burns, especially if it takes longer to regain upright posture.

### Policy

Participants articulated that governmental or institutional policies can impact the incidence and consequences of falls, calling for enforcement of accessibility laws. Many advocated for better accessibility, primarily more ramps, elevators, handrails on each side of stairs and slopes, wider curb cuts, and accessible bathrooms. Participants advocated for more seating areas in schools, public spaces, and healthcare buildings. Residents in wintery areas desired prompt snow and ice removal and salting of walkways.

### Healthcare

A need for improvements within the healthcare system was expressed, emphasizing the importance of effective therapy, treatments, and adaptive equipment across the lifespan. A common request was teaching proper falling techniques, particularly in childhood. Contrarily, others said they had been taught or naturally learned techniques, such as rotating to minimize injury. Adults highlighted lasting benefits of physical therapy on reducing fall risk, especially if it emulates real-world spaces. They desire greater access to lifetime therapy. Some participants felt healthcare professionals provided little or irrelevant fall information, especially as an adult. Lastly, conducting therapy in realistic conditions was advocated for, including simulated curbs and slopes or having therapy in less controlled spaces. Treatments (e.g., botulinum toxin, selective dorsal rhizotomy, braces, orthopedic surgeries) were perceived to help or worsen trip or fall incidence for different participants. Those living in rural spaces expressed the need for better adaptive equipment for rugged terrain.

## DISCUSSION

This study explored the causes and impacts of falls in individuals with cerebral palsy (CP) across the lifespan, highlighting the complex interplay of various factors. Many acknowledged falls as a part of life, shaping how they navigate the world and manage their fall-related concerns through various adaptation or avoidance methods to stay safe while still finding self-fulfillment. However, modifications to health-related behaviors, others’ reactions/behavior, inaccessible environments, healthcare practice, and policy are needed to systematically mitigate fall risk and repercussions.

Physical characteristics and comorbidities related to CP cause falls and change with age, based on our findings. Foot clearance is problematic. Many, including balance and fatigue, start to worsen in adulthood and are perceived to also cause functional decline.^4,16–18^ Muscles tensing in response to participants’ emotional state was reported and may represent dystonia, though it is typically underdiagnosed,^19,20^ but could increase fall risk. Epilepsy and seizures are associated with increased risk of accidents and falls in the general population^21^ and was noted here. One participant attributed their child’s fall propensity to CVI. Like dystonia, CVI is underdiagnosed but suspected to affect as many as 70% of this population.^22^ Studies are warranted to elucidate the patient characteristics that have the greatest effect on fall risk and determine effective interventions.

The direct physical consequences of falls were omnipresent. The youthful ability to “bounce back” after a fall may result in falls becoming insidious and ignored at clinical appointments in adulthood when getting up becomes more challenging. Fractures are common^3^ and concerning since adults with CP face mortality and cardiovascular disparities after fractures^23,24^ and are at a higher risk of prolonged recovery, significantly impacting independence and quality of life. Adults experience fall consequences to a greater degree due to factors such as lower bone density, reduced muscle strength, and accelerated aging.^14,25^ Individuals yearn for solutions to these problems,^18^ and participants recognized that exercise could be one solution. Empirical data on the type and incidence of fall-related injuries, recovery, and prevention interventions is needed. Such information will help justify access to physical therapy or assistive devices to support mobility for individuals with CP, since these factors were cited barriers to more proactive clinical management.

Despite the huge implication of fall-related physical injuries, one key finding was the impact of falls on emotional responses and mental health was widespread and may have a greater impact on one’s overall health than physical injuries. For the first time, panic attacks were documented in this population due to falls. Less extreme emotional responses, like worry or anxiety, can manifest as poor balance,^26^ as can mental fatigue (from concentrating on the environment and not falling) or increased cognitive load.^27^ This, in turn, increases risk of future falls. Some participants were resilient or unbothered by falls, but most participants voiced persistent concerns of anxiety, loss of confidence, loss of independence, and intentional activity avoidance, which aligns with previous studies.^3–8^ Embarrassment was the most common emotion. Because embarrassment requires one to evaluate themself against a social norm, alleviating it can be achieved by addressing one’s self-evaluation *or* addressing the social norm.

Changing social norms and the acceptance of falls can especially help adolescents and adults who may feel falls are less socially acceptable compared to when they were a child.^5^ Collectively, the findings emphasize the critical importance of asking children and adults about the emotional impact of falls and downstream effects on their life. Clinical tools like the Three Key Questions are available.^28^ Referring the patient to a mental health professional may sometimes be necessary.

Understanding the physical and psychological impacts of falls is crucial, as they can independently or collectively result in activity restriction from a young age, influencing participation in daily life and shaping long-term behaviors, place of residence, and career.^12,18^ Individuals may decide to refrain from socializing.^5,8^ This behavior can significantly impact mental health, particularly when paired with avoiding crowds or certain environments because of concern about falling or getting up. Such avoidance could indicate or lead to conditions like agoraphobia or post-traumatic stress symptoms, which are often under-recognized or underdiagnosed in individuals with CP.^29^ From a physical health perspective, persistent activity restriction (due to an injury or fear of falling) can reduce fitness and bone health, placing individuals at greater risk of serious injury and sedentary-related health conditions. Activity restriction or decreased participation may also result in cognitive issues, anxiety, and depression. The inevitable effects of aging magnify these factors, creating a vicious cycle of self-perpetuating health challenges.

Insights from our study suggest addressing fall causes and the resulting consequences requires a multifaceted approach to create physically and psychologically safer environments. Proposed solutions combine individualized strategies, healthcare support, societal changes, and improved accessibility. Individualized strategies are largely mental—resilience, adaptation, and preplanning. Others have documented similar risk management or adaptations to remain socially active.^5,8^ Some strategies are learned while others may need to be taught.

Our data highlights healthcare and clinical research’s role in developing solutions. Children may believe that they will outgrow falling, but it is a problem for all ages of ambulatory individuals.^3,9^ Therefore, access to relevant fall-related healthcare services and education across the lifespan is sorely needed.^14,12^ Safe falling strategies should be taught at all ages; this requires therapist training and access to equipment for their and their patients’ safety. Furthermore, participants suggested that therapy in less controlled or familiar spaces, including crowds and public spaces, would improve generalizability to high-risk fall situations, aligning with prior research emphasizing tailored fall prevention and the desire for healthcare professionals to understand daily challenges in CP care.^5,8,12,14,18^ Research is warranted to identify the prevalence and burden of falls, who is at greatest risk, and which interventions and supports are most effective for prevention.

From a societal perspective, individuals felt that being physically and psychologically safer involves changing the environment and public’s behavior. To enhance participation, society needs to prioritize more inclusive activities and environments, especially at schools during a child’s formative years. Inappropriate or offensive public behavior was an impediment mentioned repeatedly and agrees with previous findings that onlookers affect self-confidence and are a source of concern.^5^ Asking how to help and showing greater patience and empathy will solve many problems and reinforce individual autonomy.

Our findings suggest that addressing the shortcomings of public accessibility and policy need to be part of the multifaceted solution. Misplaced or inaccessible features were common, leading people to feel unsafe and restrict community engagement.^30^ Furthermore, unsafe or inaccessible environments can lead to absenteeism or individuals being forced out of work, resulting in significant economic and social impacts.^12^ While there were some conflicting perspectives for design specifics (e.g., more slopes vs. more low incline stairs), it was clear public accessibility needs to be enhanced. This can be achieved by including those with disabilities in the design of public spaces and a commitment from businesses to exceed the minimum accessibility requirements. Significant fear or anxiety around downslopes was highlighted here and elsewhere.^5^ Designing a shallower pitch with bilateral handrails will ensure the space is wheelchair accessible and safer for assistive device users. Standards or laws are only as effective as their enforcement; greater enforcement of disability policies, regulations, and laws are merited. Collectively, all these supportive actions foster a safe environment, mitigate the psychosocial distress of falls, and encourage participation rather than avoidance.^5,8^

### Limitations

Despite the novelty and strengths of this study, limitations remain. Participants may not represent the broader population as they were predominantly from a US Midwest hospital, ambulatory, able to participate online, their own legal decision-makers, maximum age of 76, and part of a larger fall study. Outspokenness or memorable experience are likely overrepresented. Additionally, proxy-reports from caregivers may inadequately represent their child’s experience. Prospective studies could address potential recall bias.

In conclusion, this study provides an important contribution on the holistic causes of falls and how children/their caregivers and adults with CP experience falls’ immediate and lasting physical and psychosocial effects. These findings, coupled with quantitative studies, can inform a comprehensive fall risk stratification and assessment based on an individual’s characteristics and behavior, environment, and society’s responses. Mitigating falls and their negative sequelae will truly take a village.

## Data Availability

All data produced in the present study are available upon reasonable request to the authors.

## Acknowledgments

First, we appreciate all participants who made this study possible. We would like to thank Rhonda Cady and Meghan Munger for their mentorship with the qualitative analysis. Our appreciation also extends to the CPRN for their support in participant recruitment. We gratefully acknowledge the Endowed Fund for Cerebral Palsy Treatment at Gillette Children’s for their financial support.

## REFERENCES

1 Boyer ER, Palmer M, Walt K, Georgiadis AG, Stout JL. Validation of the Gait Outcomes Assessment List questionnaire and caregiver priorities for individuals with cerebral palsy. Dev Med Child Neurol 2022; 64: 379–86.

2 Gordon AB, McMulkin ML, Baird GO. Modified Goal Attainment Scale outcomes for ambulatory children: with and without orthopedic surgery. Gait Posture 2011; 33: 77–82.

3 Esterley MT, Krach LE, Pederson K, Wandersee NG, Tierney SC, Boyer ER. Physical and psychosocial consequences of falls in ambulatory individuals with cerebral palsy by age and gross motor function. Arch Phys Med Rehabil 2024.

4 Bell BG, Shah S, Coulson N, McLaughlin J, Logan P, Luke R, Avery AJ. The impact of ageing on adults with cerebral palsy: the results of a national online survey. BJGP Open 2023; 7.

5 Gjesdal BE, Jahnsen R, Morgan P, Opheim A, Mæland S. Walking through life with cerebral palsy: reflections on daily walking by adults with cerebral palsy. International Journal of Qualitative Studies on Health and Well-being 2020; 15: 1746577.

6 Morgan P, McDonald R, McGinley J. Perceived cause, environmental factors, and consequences of falls in adults with cerebral palsy: a preliminary mixed methods study. Rehabil Res Pract 2015; 2015: 196395.

7 Morgan PE, McGinley JL. Falls, fear of falling and falls risk in adults with cerebal palsy: A pilot observational study. European Journal of Physiotherapy 2013; 15: 93–100.

8 Towns M, Lindsay S, Arbour-Nicitopoulos K, Mansfield A, Wright FV. Balance confidence and physical activity participation of independently ambulatory youth with cerebral palsy: an exploration of youths’ and parents’ perspectives. Disabil Rehabil 2022; 44: 2305–16.

9 Boyer ER, Patterson A. Gait pathology subtypes are not associated with self-reported fall frequency in children with cerebral palsy. Gait Posture 2018; 63: 189–94.

10 Morgan P, Murphy A, Opheim A, McGinley J. Gait characteristics, balance performance and falls in ambulant adults with cerebral palsy: An observational study. Gait Posture 2016; 48: 243–8.

11 Moreland B, Kakara R, Henry A. Trends in nonfatal falls and fall-related injuries among adults aged >/=65 years - United States, 2012-2018. MMWR Morb Mortal Wkly Rep 2020; 69: 875–81.

12 Shah S, Avery A, Bailey R, Bell B, Coulson N, Luke R, McLaughlin J, Logan P. The everydayness of falling: consequences and management for adults with cerebral palsy across the life course. Disabil Rehabil 2024: 1–9.

13 Organization WH. International classification of functioning, disability, and health: ICF. Geneva.

14 Cook G, Cassidy E, Kilbride C. Understanding physiotherapy and physiotherapy services: exploring the perspectives of adults living with cerebral palsy. Disabil Rehabil 2023; 45: 1389–97.

15 Gale NK, Heath G, Cameron E, Rashid S, Redwood S. Using the framework method for the analysis of qualitative data in multi-disciplinary health research. BMC Med Res Methodol 2013; 13: 117.

16 Morgan P, McGinley J. Performance of adults with cerebral palsy related to falls, balance and function: a preliminary report. Dev Neurorehabil 2013; 16: 113–20.

17 Gannotti ME, Sarmiento CA, Gross PH, Thorpe DE, Hurvitz EA, Noritz GH, Horn SD, Msall ME, Chambers HG, Krach LE. Adults with cerebral palsy and functional decline: A cross-sectional analysis of patient-reported outcomes from a novel North American registry. Disabil Health J 2024; 17: 101593.

18 Cohen J. Family reflections: a whole person: navigating aging and cerebral palsy. Pediatr Res 2025.

19 Gilbert LA, Gandham S, Ueda K, Chintalapati K, Pearson T, Aravamuthan BR. Upper Extremity Dystonia Features in People With Spastic Cerebral Palsy. Neurol Clin Pract 2023; 13: e200207.

20 Aravamuthan B, Pearson TS, Chintalapati K, Ueda K. Under-recognition of leg dystonia in people with cerebral palsy. Ann Child Neurol Soc 2023; 1: 162-7.

21 Mahler B, Carlsson S, Andersson T, Tomson T. Risk for injuries and accidents in epilepsy: A prospective population-based cohort study. Neurology 2018; 90: e779–e89.

22 Fazzi E, Signorini SG, R LAP, Bertone C, Misefari W, Galli J, Balottin U, Bianchi PE. Neuro-ophthalmological disorders in cerebral palsy: ophthalmological, oculomotor, and visual aspects. Dev Med Child Neurol 2012; 54: 730–6.

23 Whitney DG, Bell S, Etter JP, Prisby RD. The cardiovascular disease burden of non-traumatic fractures for adults with and without cerebral palsy. Bone 2020; 136: 115376.

24 Whitney DG, Bell S, Hurvitz EA, Peterson MD, Caird MS, Jepsen KJ. The mortality burden of non-trauma fracture for adults with cerebral palsy. Bone Rep 2020; 13: 100725.

25 Whitney DG, Hurvitz EA, Caird MS. Critical periods of bone health across the lifespan for individuals with cerebral palsy: Informing clinical guidelines for fracture prevention and monitoring. Bone 2021; 150: 116009.

26 Hall KJ, Van Ooteghem K, McIlroy WE. Emotional state as a modulator of autonomic and somatic nervous system activity in postural control: a review. Front Neurol 2023; 14: 1188799.

27 Salihu AT, Hill KD, Jaberzadeh S. Age and Type of Task-Based Impact of Mental Fatigue on Balance: Systematic Review and Meta-Analysis. J Mot Behav 2024; 56: 373–91.

28 Prevention CfDCa. Pocket Guide Preventing Falls in Older Patients. In: Control NCfIPa editor.; 2019.

29 Bhatnagar S, Mitelpunkt A, Rizzo JJ, Zhang N, Guzman T, Schuetter R, Vargus-Adams J, Bailes AF, Greve K, Gerstle M, Pedapati E, Aronow B, Kurowski BG. Mental Health Diagnoses Risk Among Children and Young Adults With Cerebral Palsy, Chronic Conditions, or Typical Development. JAMA Netw Open 2024; 7: e2422202.

30 Bonehill J, Von Benzon N, Shaw J. ‘The shops were only made for people who could walk’: impairment, barriers and autonomy in the mobility of adults with Cerebral Palsy in urban England. Mobilities 2020; 15: 341–61.

